# Computational modeling of the temporal influences between cues, craving and use in addiction: A dynamical system analysis based on ecological momentary assessment data

**DOI:** 10.1101/2025.01.13.25320446

**Authors:** Christophe Gauld, Damien Depannemaecker, Marc Auriacombe, Fuschia Serre

## Abstract

Substance Use Disorders (SUD) can be conceptualized as a prospective link from cues to craving and use. To explore the nonlinear relationships between craving and cues, this study applied dynamical systems theory (DST) to ecological momentary assessment (EMA) data. Optimized linear Seasonal Auto-Regressive Integrated Moving Average with eXogenous variable (SARIMAX) models were used to phenotype patients with SUD (alcohol, tobacco, cannabis, opiates, and cocaine), considering the potential for complex interactions between cue exposure and craving intensity in daily life. These phenotypic profiles were replicated in computational DST models to analyze the nonlinear interactions between cues, craving, and use. The study involved 211 individuals and 8,260 observations, with 154 patients fitting the SARIMAX model for the influence of cues on craving, and 57 patients fitting the SARIMAX model for a possible influence of craving on cues. Two DST models were adjusted to replicate the complex temporal dynamics of SUD based on these two directions of influence. The first DST model (adjusted to the influence of cues on craving) showed that an increase in cues leads to a rise in craving, which then diminishes both cues and craving itself, with use patterns following craving’s trajectory. This patient profile is driven by a phenomenon of “maximum cue saturation”. The second DST model (adjusted to the influence of craving on cues) demonstrated that an increase in craving was followed by an increase in cue reporting, leading to use, with use peaking and then reducing craving. This patient profile is characterized by a phenomenon of “maximum use saturation”. Both models highlight craving as an essential modulator between cues and use, opening new therapeutic avenues.

## Introduction

Substance Use Disorder (SUD) is characterized by recurrent relapses despite efforts to reduce use or abstain [1]. One of the main objectives of SUD treatment is to reduce the frequency of relapses over time. As an important symptom of SUD [1–8], the fluctuating phenomenon of craving can be captured by daily-life monitoring systems such as ecological momentary assessment (EMA) [9]. EMA, adapted and validated to study craving in SUD [10], allows a real-time measurement of individual momentary states in the natural environment through repeated assessments overtime. Based on EMA research, a human model of addiction has been suggested across several substances and behaviors: the cues – craving – use model [1, 4, 11]. In this model, an increase in the number of reported cues is associated with an increase in the intensity of self-reported craving in the following EMA survey, itself associated with an increased likelihood of use in the following EMA survey. This craving – use link has been considered unidirectional [11–14] using mixed models like hierarchical linear models (HLM), which allows for the analysis of data with a nested structure, accounting for variability at multiple levels of observation. However, HLM has some limitations such as the difficulty in accounting for cross-effects, i.e., integrating complex interactions between levels. Furthermore, they are only linear: the effects (e.g., craving) may be directly proportional to the causes (e.g., cues) – as opposed to non-linear models where the relationships between variables can be disproportionate, interactive, producing effects which are not predictable simply by addition or multiplication. Finally, HLM cannot account for multidirectionality, e.g., that cues and craving could influence each other in a loop. Due to the important intra-individual variability and its intrinsic non-linearity, the evolutionary dynamics of a specific patient with a SUD are particularly complex to model.

To go further, modeling the evolution of SUD variables and their interactions requires finding alternatives to traditional statistical methods [2]. A growing literature based on dynamical systems theory (DST) seeks to capture the temporal dynamics of psychiatric conditions (e.g., [15–35]), including SUD. DST accounts for the evolution of clinical manifestations over time without abolishing the complexity of the relationships between variables and their multidirectionality, allowing the prediction of different types of temporal evolution. This theory is based on dynamic objects (e.g., tipping points, transitions, steady states, strange attractors, limit cycles or oscillations) and dynamic concepts (e.g., emergence, instability, or variations related to fluctuations in initial conditions – referring to the “butterfly effect”, etc.). However, most psychiatric studies include biological factors (e.g., [36]) and few studies have empirically included psychiatric clinical manifestations (e.g., [19]).

Based on EMA data, considering the possibility that, in addition to a unidirectional prospective link from cues to craving, an increase in craving could change perception of cues [37], DST models allow the representation of dynamics and mutual influences between variables. The main goal of this study was to model alternative evolutions of craving and cues, to explore their influence on substance use. Such results could lead to proposing a classification of individuals with SUD based on dynamics’ profiles, as well as providing information on the attractive states (i.e., the states to which the system gravitates) – allowing to understand what stabilizes the patient in a given (healthy or pathological) state.

## Methods

This study is based on two successive stages: i) the use of optimized linear statistical models (SARIMAX) to identify, based on EMA data, two patient profiles according to the direction of influence between cues and craving; ii) the use of a computational model (DST) to analyze the complex relationships between cues and craving according to these profiles. In line with the classic cues-craving-use model (in which an increase in cues can linearly contribute to an increase in craving), we hypothesized that a state of craving could make an individual more “sensitive” or more “receptive” to cues, with an effect on use, creating a feedback loop: cues lead to craving, which increases cue perception, leading to more craving, and ultimately, more use. This reflects a non-linear dynamic where continuous mutual interactions bring about a dynamic process. The modeling focuses on the cues-craving relationship, as use is the outcome.

### Data collection in EMA

#### Participants

Data were extracted from two EMA studies with similar protocol [10, 43], which comprised individuals aged 18 years and above, who were beginning addiction outpatient treatment (in Bordeaux and Bayonne, France), were diagnosed with at least one DSM-5 Substance Use Disorder for tobacco, alcohol, cannabis, cocaine or opiates, and were free from active psychosis or severe cognitive impairment [38]. Informed consent was obtained, and the studies were approved by the French Regulation and ethical committee (CPP SOOM III/DC-2009/01; CNIL/DR-2015-408; CPP IDF X: 11-2019/IR-RCB: 2018-A00952-53; CNIL/MR003). All methods were performed in accordance with the relevant guidelines and regulations. Participants received standard comprehensive care, consisting of individual behavioral treatment focused on relapse prevention and psychosocial support combined, when available, with pharmacotherapy. After establishing a target quit date according to the patient’s personal goals, full abstinence or use reduction was encouraged as an outcome, but with no negative consequences for the patient if failure to achieve this goal.

#### Assessments

Each participant underwent an initial interview to collect sociodemographic variables and substance use-related information using a validated French version of the Addiction Severity Index (ASI) [39, 40], modified to consider tobacco addiction. The Mini International Neuropsychiatric Interview–Plus (MINI) structured interview [41] was adapted to evaluate the current DSM-5 SUD status within the past year. When multiple SUD co-occurred, the primary substance was determined according to the main problematic substance reported by the individual, and on which the treatment was focused.

#### Ecological momentary assessment

The EMA protocol was previously described in detail [10]. EMA assessments began within the first month after treatment initiation. Following a training session, each participant received a tablet to carry for 14 days. Each tablet was programmed to administer four electronic surveys per day, inquiring about primary substance use, craving, and the presence of cues. The signal schedule was adjusted to accommodate participants’ usual sleep patterns. Financial compensation was provided based on the number of surveys completed, with a maximum of 100 euros for participants who completed 75% or more of the assessments. Methodological details and comparisons with other studies using this validated protocol [11, 42–44] are presented in **Supplementary Material 1**.

#### Data of interest

At each electronic assessment, participants were asked to rate the maximum intensity level of desire to use their primary substance they had felt since the previous assessment on a 7-point scale (1 = no desire to use 7 = extreme desire to use). This item was referred to as “craving”. Participants were also asked if they had encountered cues related to their primary substance, selected from a personalized list for each individual. This item was labeled “cues” and included specific objects or contexts typically associated with substance use, such as materials used for consumption (e.g., a bottle), the sight or smell of the substance, or places associated with use (e.g., a bar). It also encompassed personal cues, identified during a personal interview, that referenced specific circumstances, objects, individuals, emotions, or environmental contexts linked to their substance use (e.g., “being with my friend Sam”) [11]. The “cues” variable is a count measure that ranges from 0 upwards, with no upper limit, reflecting the total number of cues reported since the previous assessment. Although these refer to “cue perceptions”, for reading convenience, we used the term “cues” below. Finally, participants were asked if they had used the primary substance, with this variable being binary (0 = no, 1 = yes), focusing solely on whether substance use occurred since the prior assessment. This item was referred to as “use”. All three questions inquired about participants’ experiences since the previous assessment.

Due to a restricted number of assessments occurring on the starting and ending day of the study, and their overlap with investigator contact, only data collected between days 2 and 13 were analyzed (n=12 days) (**Table 2**).

**Table 1.** Characteristics of the sample (n=211 participants): primary substance, demographic variables, substance use-related information and psychiatric comorbidity. ASI: Addiction Severity Index; MINI: Mini International Neuropsychiatric Interview–Plus; SD: standard deviation.

|  | Sample characteristics | All participants (n=211) |  |
| --- | --- | --- | --- |
|  |  | % (n) | Mean (SD) |
| Primary substance | Alcohol | 35 (73) | – |
|  | Tobacco | 28 (58) | – |
|  | Cannabis | 18 (39) | – |
|  | Opiates | 16 (34) | – |
|  | Cocaine | 3 (7) | – |
| Demographic variables | Sex (female) | 36 (78) | – |
|  | Employed | 56 (116) | – |
|  | Age | – | 38.2 (11.4) |
|  | Education (years) | – | 12.5 (2.74) |
|  | Pharmacological treatment | 79 (167) | – |
| Substance use-related information and psychiatric comorbidity | ASI addiction severity (0-9) | – | 6.6 (0.76) |
|  | Mood disorder on the MINI (current diagnosis) | 34 (72) | – |
|  | Anxiety disorder on the MINI (current diagnosis) | 45 (93) | – |

**Table 2.**
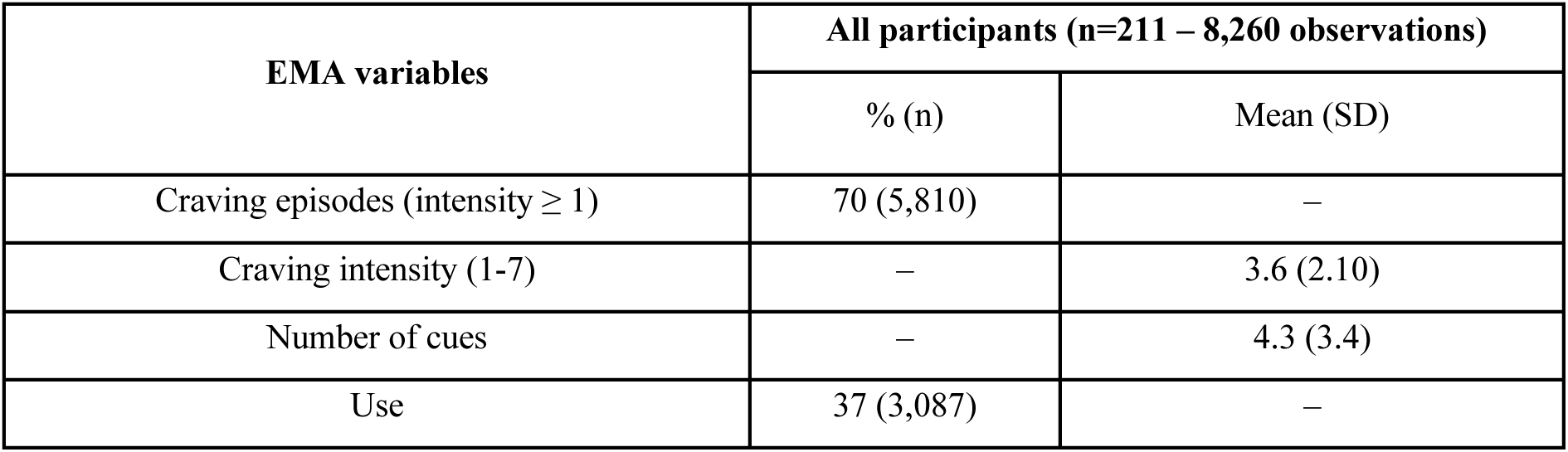
Ecological Momentary Assessment (EMA) report of craving episodes and intensity, number of cues, and reports of primary substance use of participants (n=211). SD: standard deviation.

### Statistical and computational models

#### Aims of the models

First, as detailed in **Supplementary Material 2**, we used optimized linear SARIMAX models with craving and cues to determine the mutual influence profiles of patients. We aimed to identify: i) a *SARIMAX-1* model, referring to the influence of cues on craving, hereinafter named “cues-to-craving” model; ii) a *SARIMAX-2* model, referring to the influence of craving on cues, hereinafter named “craving-to-cues” model.

Secondly, we used the SARIMAX model 1 to determine the specific parameter values of a model based on the DST (*DST-model 1*), and the SARIMAX model 2 to determine the specific parameter values of another model based on the DST (*DST-model 2*). We qualitatively observed the complex evolution of each of the two variables within the two DST models. “Use” was treated as an output of the model, with its values deliberately left unconstrained to follow the parameterized dynamics of the cues-craving structure in both DST models. This approach allowed us to capture the intricate interactions between cues, craving, and use.

By combining linear SARIMAX models with DST models, we aimed to create a framework that captures both the linear and nonlinear dynamics of craving and cues. The SARIMAX models first classified patients based on their interaction profiles. These profiles served to establish parameter values for the DST models, which further analyze the nonlinear interactions between craving and cues. This approach allowed us to utilize linear models for the initial sequence of variables without necessarily providing a parameter characterization. Subsequently, the DST models determine these parameters to capture intricate feedback loops between craving, cues, and use. Though SARIMAX models were constrained by assumptions of linearity and stationarity, this hybrid approach could offer an interesting tool for understanding complex craving dynamics across varied profiles. As we will describe later, the comparison of SARIMAX and DST models is a methodological challenge due to differences in scope and (non)linearity. Specifically, in the DST framework, the parameter ranges are reconstructed to fit the data, rather than being directly calculated from empirical measurements. This approach emphasizes conceptual alignment with circadian-scale dynamics rather than exact value matching.

To ensure the reliability of our method, we conducted a simulation study by generating two groups based on distinct craving-to-cues and cues-to-craving dynamics, according to the DST model parameter distributions. We then verified our ability to recover these groups using SARIMAX modeling, applying separate models to identify each directional influence. Finally, based on the bifurcation analysis described below, we checked whether the parameter values of the DST model could be recovered for each of the two groups, ensuring consistency between the simulated data and the underlying DST model parameters (**Supplementary Material 3**).

#### SARIMAX models

SARIMAX models, extensively detailed in **Supplementary Material 2**, are linear algorithms for forecasting data, considering past values (AR – AutoRegressive) and data noise (MA – Moving Average) to predict future values [45].

##### Stationarity and seasonality assessments of the EMA data

Stationarity is a condition of validity for this time-series analysis using the Auto-Regressive Moving Average (AR-MA) method. The stationarity of the time-series data for craving and cues was assessed using the Augmented Dickey-Fuller (ADF) test [46]. If the time series was not stationary, an integration factor (I(*d*)) was added to the model, allowing differentiation to make the data stationary (i.e., resulting in a “AR-I-MA” model). Seasonality (classically denoted *S* in the model, providing a “S-ARIMA” model) was considered given the repeated daily nature of the data. Finally, an exogenous variable (denoted *X* in the model, resulting in a “SARIMA-X” model) could be added to model an additional effect on the time series of the first variable (i.e., cues for the craving time series, and craving for the cues time series) [47, 48].

##### Model fitting

For each patient, two SARIMAX models were fit to the data: one where craving was modeled as the dependent variable influenced by cues (*SARIMAX-model 1* – “cues-to-craving” model), and another where cues were the dependent variable influenced by craving (*SARIMAX-model 2* – “craving-to-cues” model). Different combinations of autoregressive and moving average terms were tested for each patient. Only the parameter values for the equations governing *y* (craving) and *z* (cues) were adjusted to align with each of the two SARIMAX models. The parameter values for the equation of *x* (use) were deliberately left unconstrained, allowing to observe its dynamics as an output variable within the models.

##### Parameter optimization

The *y* and *z* parameters of the two SARIMAX models were optimized individually for each patient. A grid search was performed across potential values of the AR and MA component, identifying the combination that minimized the Akaike Information Criterion (AIC). This criterion balances model fit and complexity to prevent overfitting. The choice to use the AIC over the Bayesian Information Criterion (BIC) was motivated by the objective of optimizing predictive accuracy while accommodating the complexity of individual temporal dynamics, as AIC imposes a less stringent penalty on model parameters, thus enhancing the model’s sensitivity to subtle within-subject variations. Two AIC were provided per patient (one for each SARIMAX model of a patient).

##### Model selection

For each patient, the preferred SARIMAX model (either *SARIMAX-model 1* or *SARIMAX-model 2*) was determined based on the lower AIC. We assume that each patient aligns with only one specific model, excluding the other. Following the identification of which participants best fit each model, we compared the two selected subgroups based on socio-demographic characteristics, substance types, addiction severity, psychiatric diagnoses, and EMA variable characteristics (**Table 3**). This comparison aimed to 1) clinically describe the profiles of patients displaying different dynamic patterns (cues to craving versus craving to cues), and 2) methodologically confirm the absence of significant differences between the groups, ensuring that any observed distinctions in model dynamics were not attributable to these underlying characteristics. Details on these models are provided in **Supplementary Material 2**.

**Table 3.**
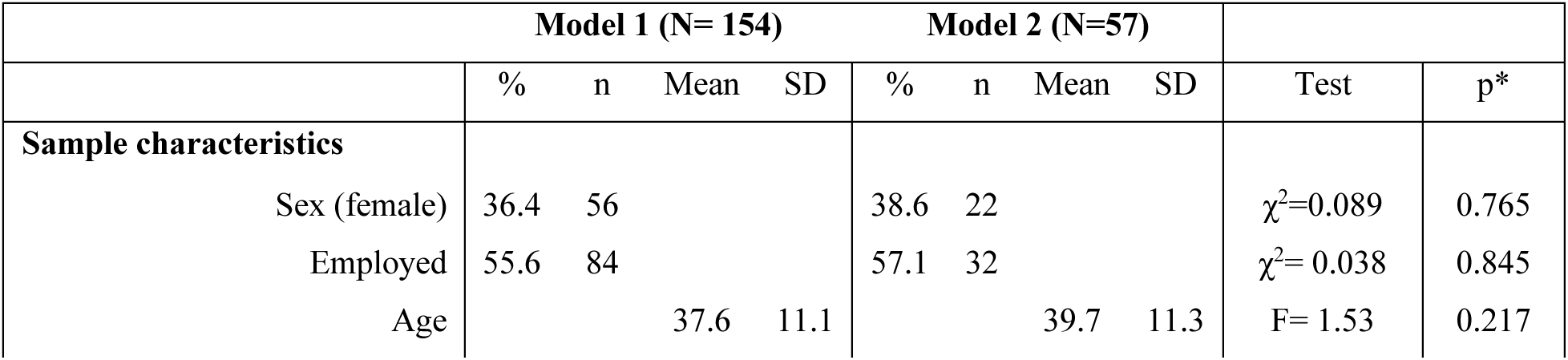
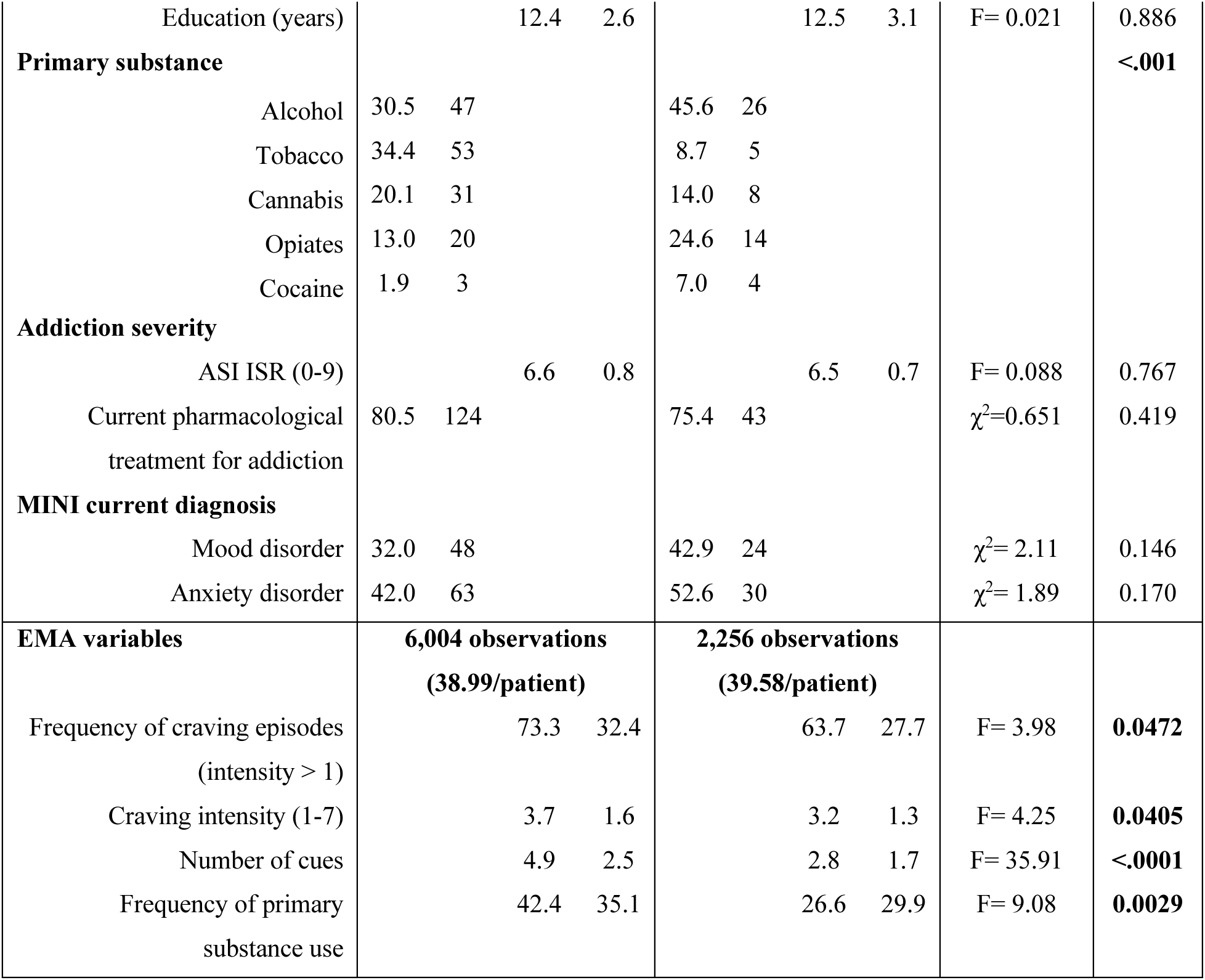
Socio-demographic characteristics, substance types, addiction severity, psychiatric diagnoses and EMA variable characteristics of the two temporal profile (SARIMAX) of patients with Substance Use Disorder (SUD) (n=211). SARIMAX: Seasonal Auto-Regressive Integrated Moving Average with eXogenous variable. “Model 1”: SARIMAX-model 1 (“cues-to-craving”); “Model 2”: SARIMAX-model 2 (“craving-to-cues”).

##### Sensitivity analysis

We then performed a sensitivity analysis on the sub-datasets of alcohol and cocaine to assess the stability of the models across these substance-specific data.

##### Auto-correlation analysis

Finally, we proposed studying the temporal dynamics between craving and cues through auto-correlation and cross-correlation analyses, allowing to identify individual temporal structures, key time lags, and descriptive statistics revealing variability in craving and cues across the sample.

#### DST models

##### Dynamical system modeling

A set of differential equations was used to model the dynamical relationship between craving and cues. The model was integrated over the 12-day observation window and extended to a theoretically infinite duration to visualize the temporal interaction of one variable on the other.

##### Details of the DST model

The detail of the DST model has been described elsewhere [28]. In a nutshell, it is based on four differential equations, producing a nonlinear representation of participant’s internal factors (i.e., subjective phenomenological experiences and/or biological elements) (*y*) and of environmental noise (*z*). Based on the empirical data collected in EMA regarding SUD, the “patient’s internal factor” corresponds to the intensity of craving (*y*), and the “environmental noise” corresponds cues (more precisely, to the reporting of cues) (*z*). Two other variables allow modulating: i) the long-term temporality of patients’ conditions (*f*); ii) and the intensity of behavior (*x*), corresponding here to substance use (passively evolving according to the two cues-craving models). Thirteen parameters are described within these four variables, with their clinical translation, presented in **Supplementary Material 4**. Details of interactions between parameters within each variable are described elsewhere [28], but the four differential equations and the thirteen parameters of the model are presented below^1^:

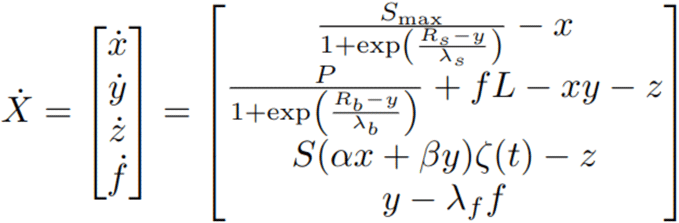

As an example, **Figure 1** presents two simulations of the DST model with different parameter values, by considering three random variables. We observe that the simulated patient trajectories in the model’s “relief” differ when the parameter values are varied.

**Figure 1.**
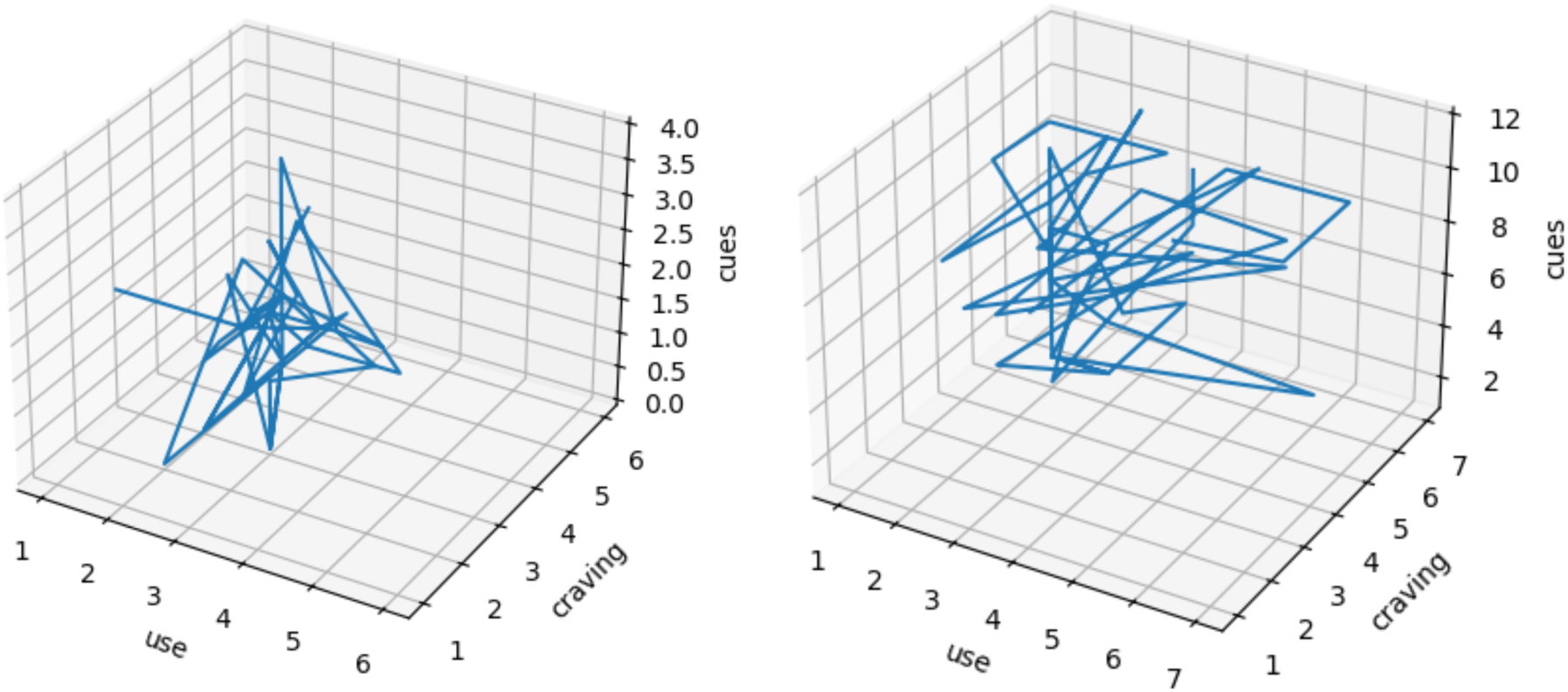
Phase space representation of the co-evolution of the cues, craving and use for two randomly chosen patient from the dataset. The relations between these variables are complex and no trivial linear interdependence appear.

##### Construction of the two DST models

We built two DST models with specific parameter values. Given the short time scale considered (tens of days), we fixed the *f* variable – originally representing predisposing factors over months and years. The parameter values in the *y* (craving) and *z* (cues) equations were adjusted to align with each of the two SARIMAX models.

To identify the regions of interest in the parameter space, we performed a bifurcation analysis. Bifurcation analysis is a well-known method in the field of dynamical systems to study how the qualitative behavior of a system changes as parameters are varied, identifying transitions such as the emergence of cycles, stability, or new equilibrium states [49, 50]. We focused on the parameters associated with the dynamics of the internal state (craving), specifically *λ*_b_, *R*_b_, and *L*. This allowed to determine the ranges of values that enable the existence of a limit cycle, corresponding to the observed circadian oscillations. Similarly, we identified the regions of the parameter space that allow for the existence of a stable fixed point, representing a situation where the craving variable does not oscillate spontaneously but remains sensitive to perturbations induced by external inputs related to cues.

To explain it differently, we identified a subset of the parameter space using bifurcation analysis, representing the possible dynamical repertoire that matches the circadian evolution (i.e., changes observed over a 24-hour day-night cycle) observed in the data. This selection was based on the features associated with temporality and rhythmicity, central to interpreting model comparisons. The parameter adjustment process relied on an iterative hands-on fit, enabling the reproduction of sequential patterns of emergence and fluctuation between craving and cues in each model. This empirical manual exploration of the parameters of the system, to align with the perceived temporal dynamics, was conducted without automated method (e.g., simulation-based inference, to be applied in future works). The simulations shown here were chosen as the best matching observation within the range identified previously.

Then, for each subject, the best-fitting linear SARIMAX model (either cues-to-craving or craving-to-cues) was selected and subsequently used to fit a nonlinear DST model, parameterized to capture their dynamics. The parameter adjustment process was conducted through successive simulations, allowing to iteratively refine the model to obtain a qualitative understanding of the system’s behavior while ensuring it aligned with the observed dynamics in the SARIMAX models. For instance, the parameter *L* (representing the “vulnerability level” in the *y* equation) may need to be set high to reflect the increasing influence of craving on cues. Such parameterization of vulnerability would constitute an interesting result: for craving to affect sensitivity to cues, a patient must have a high level of vulnerability (e.g., genetic predispositions, psychological vulnerability, neurological comorbidities, etc.).

All the analyzes were performed with Python [51].

### Code availability

The code is added to the open access repository: https://github.com/ChristopheGauld/Dynamical_systems_for_craving.

## Results

### Sample description and assessments

As described in **Table 1**, 211 individuals were included in the study. The mean age was 38.2 years old and 36% were female (n=36). Primary substance was: 35% alcohol, 28% tobacco, 18% cannabis, 16% opiates and 3% cocaine. The flow-chart of participant inclusion is presented in **Supplementary Material 5**.

### EMA assessments

Among the 10,128 EMA electronic assessments planned and delivered, 8,260 (82%) were completed. The majority of EMA assessments reported episodes of craving (70% with non-null intensity), and 37% reported primary substance use (**Table 2)**.

### SARIMAX models

The ADF test confirmed that the time-series does not have a time-dependent structure (p = 0.0003 for craving and p << 0.001 for cues, indicating that both series are stationary), supporting stationarity for each patient and for each variable (cues, craving and use). No detrending was necessary. No seasonality, which could have been present here due to typical day-by-day fluctuations, was found in the data. SARIMAX models could thus have been named “ARMAX”, but we keep the initial term to ensure understanding.

After testing the two SARIMAX models for each patient and after individual optimization, we found 154 patients who fit better (according to the AIC) with the SARIMAX-model 1 (“cues-to-craving”) and 57 patients who fit better with the SARIMAX-model 2 (“craving-to-cues”). As expected in the analysis seeking to maximize the fit of a patient with one of the two models, no patient belonged to both models. Details on the AIC of the 1 and 2 SARIMAX models and on the preferred model according to the lowest AIC are given in **Supplementary Material 2**. **Table 3** provides the characteristics of the patients who best fit each of the two SARIMAX models. There were no significant differences in the distribution of sociodemographic characteristics, addiction severity, psychiatric diagnoses, or substance types between patients fitting each model; however there were differences for some of the EMA variables (see **Table 3**).

The results of the sensitivity analysis and of the auto-correlation analysis between craving and cues are presented in **Supplementary Material 2**.

### DST models

Using successive simulations based on different parameterization of the *z* (cues) and *y* (craving) variables of the DST model, we sought to represent in a non-linear way (unlike SARIMAX models) the mutual interactions between craving and cues. We did not seek to modify the other two variables (*f* variable for long-term temporality or *x* variable for use). The parameter values of the equations of *y* (craving) and *z* (cues) equations are modified in order to fit with each of the two SARIMAX models. We defined two sets of parameters: the first set of parameters of the *DST-model 1,* corresponds to the dynamical system representation of the 154 patients who fit with the *SARIMAX-model 1*; the second set of parameters of the *DST-model 2,* corresponds to the dynamical system representation of the 57 patients who fit with the *SARIMAX-model 2*.

The details of the parameter values of the two DST models are provided in **Table 4**. Five parameter values were necessarily modified to recreate the two SARIMAX-models: *L, R_b_*, *P, τ_x_*, and *τ_y_* (in red in **Table 4**). Firstly, the value of the *L* parameter (corresponding to the vulnerability level in the *y* equation) was barely modified (*L* = 1.01 in the DST-model 1 and *L* = 1.00 in the DST-model 2) to obtain these two profiles of influence of craving on cues. Secondly, the value of the parameter *R_b_* (referring to the sensitivity of the internal elements) was necessarily modified (*R_b_* = 1.04 in the DST-model 1 and *R_b_* = 0.904 in the DST-model 2): for craving to have an influence on sensitivity to cues (DST-model 2), a patient must have a lower level of sensitivity. Thirdly, the value of the parameter *P* (representing potentiation, or the maximal rate of internal elements amplifying symptoms) was necessarily increased for DST-model 1 (*P* = 10) compared to DST-model 2 (*P* = 1), indicating that craving (which drives DST-model 1) serves as an internal mechanism that amplifies use. Finally, τ_x_ and τ_y_ refer solely to the time scales specific to each variable (e.g., τ_x_ • *dx/dt*) without clinical interpretation, and their increase in DST-model 1 does not alter the interpretation of the other parameter values.

**Table 4.** Parameter values of the two dynamical system models fitted with linear models (n=211). DST: Dynamical System Theory. The details of the parameters and their clinical translation are given in Supplementary Material 4.

| DST models | Parameters of x, y and z variables |  |  |  |  |  |  |  |  |  |  |  |  |  |
| --- | --- | --- | --- | --- | --- | --- | --- | --- | --- | --- | --- | --- | --- | --- |
|  | x |  |  |  | y |  |  |  |  |  | z |  |  |  |
|  | S<sub>max</sub> | R<sub>s</sub> | λ<sub>s</sub> | τ<sub>x</sub> | B<sub>max</sub> | R<sub>b</sub> | λ<sub>b</sub> | P | L | τ<sub>y</sub> | S | α | β | τ<sub>z</sub> |
| *DST-model 1* | 10 | 1 | 0.1 | 14 | 10 | 1.04 | 0.05 | 10 | 1.01 | 14 | 1.0 | 0.5 | 0.5 | 1 |
| *DST-model 2* | 10 | 1 | 0.1 | 2 | 10 | 0.904 | 0.05 | 1 | 1.00 | 2 | 1.0 | 0.5 | 0.5 | 1 |

Dynamics of craving and cues on the *DST-model 1* and on the *DST-model 2* are represented in **Figure 2** and **Figure 3**, respectively. Within these two figures, although there was no fixed scale due to the nonlinear interactions between relative dynamics, it was important to consider that the baseline represented an absence of interaction, visualized by an increase in a variable (e.g., the baseline for craving represented the level corresponding to the absence of craving, when it did not spontaneously increase or respond to another variable).

**Figure 2.**
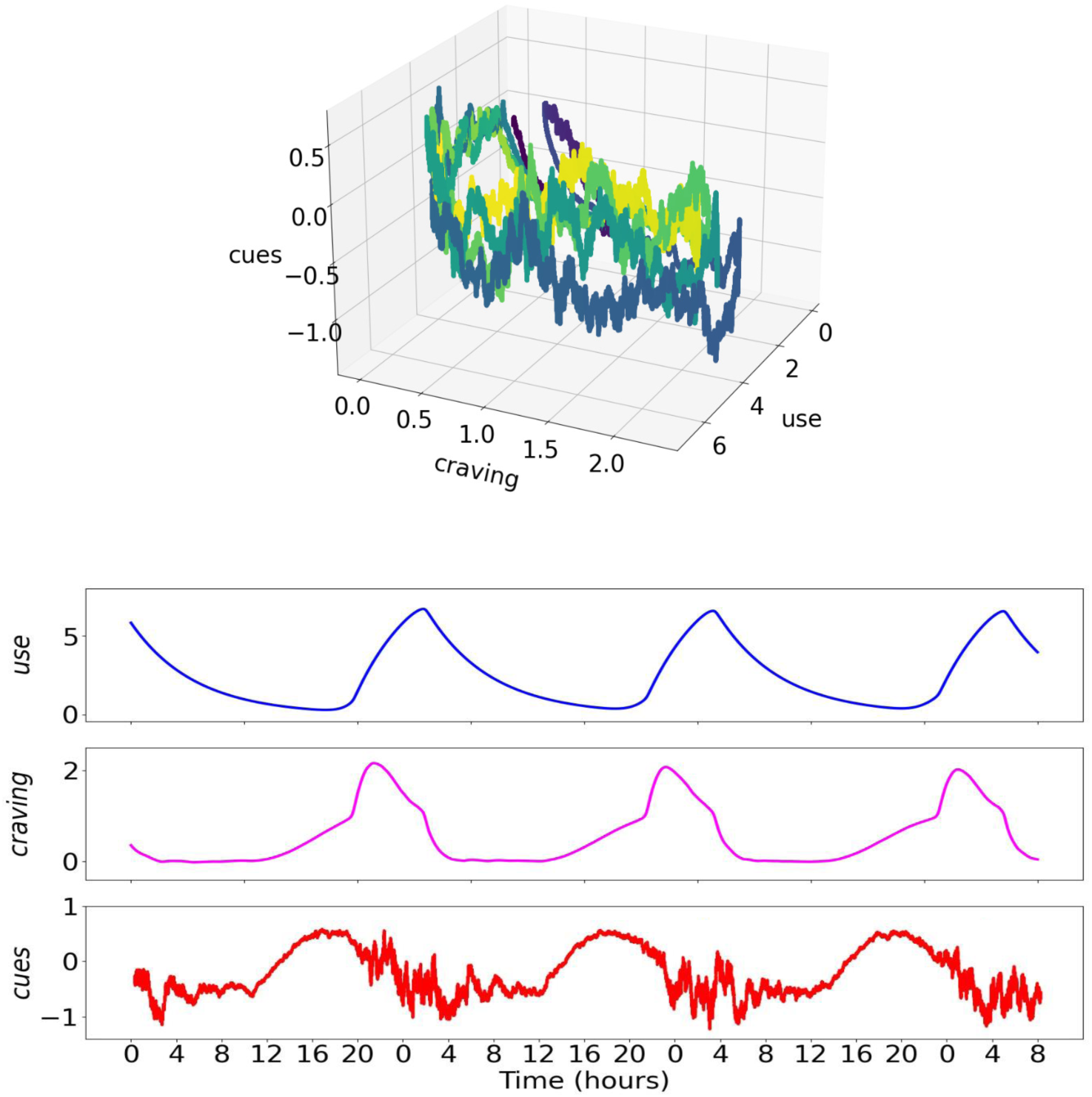
Dynamic representation of the temporal profile of patients with Substance Use Disorder (SUD) belonging to the *SARIMAX-model 1* (“cues-to-craving”) (n=154), for 4 days. **Top**: Space phase of the model, i.e., the conceptual framework illustrating all possible states of the system and the interactions between variables over time. This 3D representation illustrates the dynamic interactions between craving, cues, and use, providing a visual model of how fluctuations in one variable correspond with variations in the others. Although such a depiction may not be immediately readable, it remains essential for capturing the complex, nonlinear relationships inherent in dynamic systems and is commonly used to elucidate interdependencies across multiple variables. **Bottom**: Temporal representation of the model (4 times 12 hours, hours are noted every 4 hours and over 24 hours for the sake of clarity). SARIMAX: Seasonal Auto-Regressive Integrated Moving Average with eXogenous variable; DST: Dynamical System Theory.

**Figure 3.**
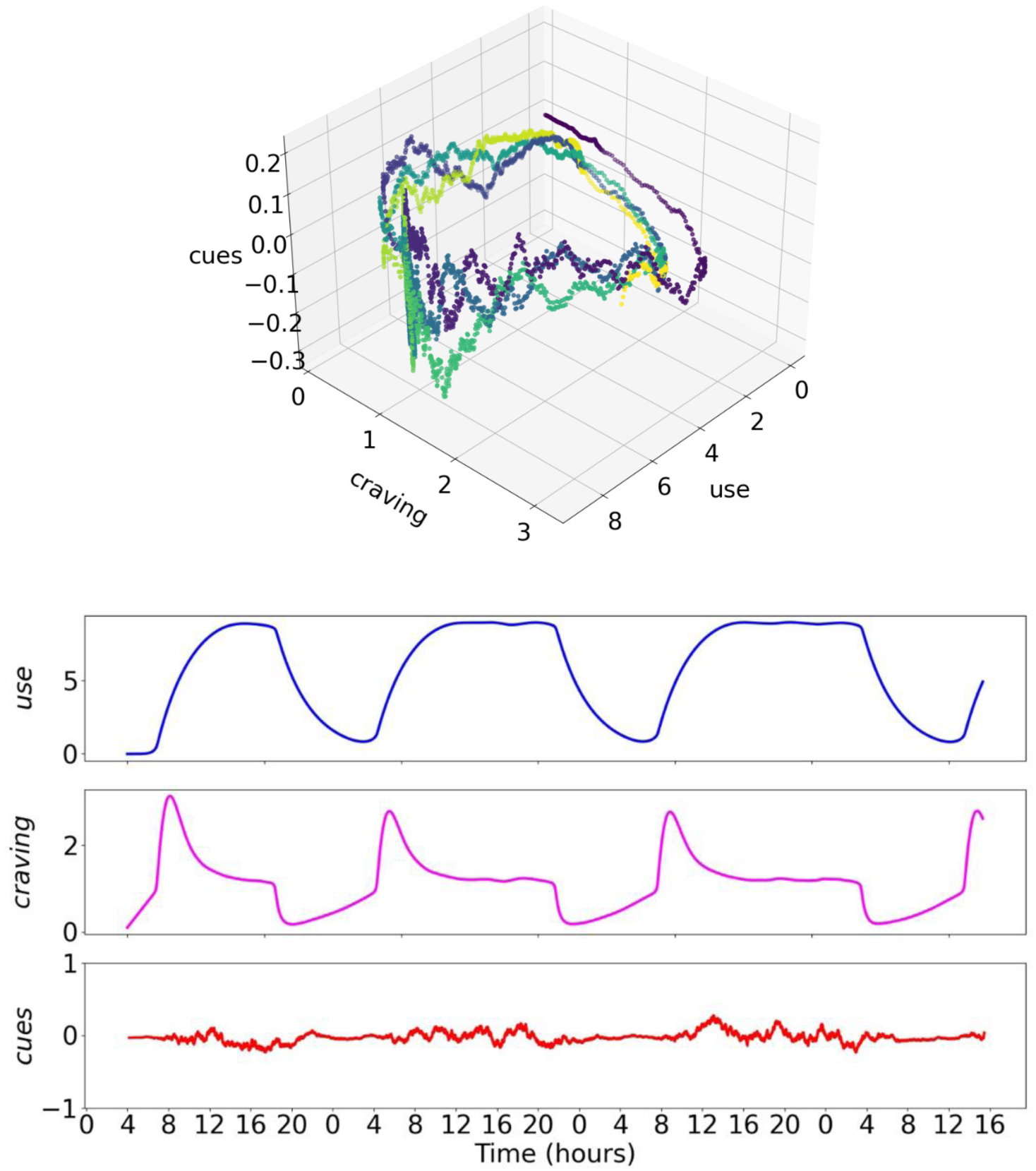
Dynamic representation of the temporal profile of patients with Substance Use Disorder (SUD) belonging to the *SARIMAX-2* model (“craving-to-cues”) (n=211), for 4 days. **Top**: Space phase of the model, i.e., the conceptual framework illustrating all possible states of the system and the interactions between variables over time. This 3D representation illustrates the dynamic interactions between craving, cues, and use, providing a visual model of how fluctuations in one variable correspond with variations in the others. Although such a depiction may not be immediately readable, it remains essential for capturing the complex, nonlinear relationships inherent in dynamic systems and is commonly used to elucidate interdependencies across multiple variables. **Bottom**: Temporal representation of the model (4 times 12 hours, hours are noted every 4 hours and over 24 hours for the sake of clarity). SARIMAX: Seasonal Auto-Regressive Integrated Moving Average with eXogenous variable; DST: Dynamical System Theory.

In **Figure 2** (DST-model 1 for *SARIMAX-model 1* patients, N=154), when an increase in cues (visible as positive events in the *z* variable) preceded craving, there was a relatively rapid increase in craving, which in turn reduced the cues, followed by a slower decrease in craving (**Figure 2**, bottom). Craving peaked in alignment with peak cues. Craving only returned to its baseline level after cues also returned to baseline. Use progressively increased throughout the duration of the craving rise (e.g., from 8 p.m. to midnight on the 1^st^ day) and only began to decrease in temporal conjunction with the relatively sudden drop in craving. This temporal overlap between the decreases in craving and use limited our ability to determine the direction of influence between the two variables. However, it is noteworthy that the decrease in reporting of cues preceded this joint decrease in craving and use (e.g., at 4 p.m. on the 1st day). The return to baseline may therefore be attributed to cue saturation, with a craving modulation (highlighted by the observation that the return to baseline occurred only after a reduction in craving).

In **Figure 3** (*DST-model 2* for *SARIMAX-model 2* patients, N=57), when an increase in craving occurred before cues, this prior craving increase influenced cue reporting, leading to greater simultaneity between the two variables compared to the previous model (**Figure 3**, bottom). Craving decreased during the period of noisy cue reporting, with both craving and cues returning to baseline simultaneously. Use increased gradually throughout the duration of the craving increase and, as before, only began to decrease with the relatively sudden drop in craving. Cue reporting returned to baseline in temporal conjunction with this decrease in use and craving (unlike the earlier return seen in the previous model). The return to the baseline state was therefore due to saturation of use, implying a modulation of craving.

Finally, the simulation study demonstrates the robustness of our modeling approach in reliably distinguishing the craving-to-cues and cues-to-craving groups, confirming the SARIMAX method’s effectiveness in capturing these distinct directional influences based on different parameter distributions of the DST models. Additionally, for each SARIMAX model, we verified the alignment of the recovered simulations with the dynamics initially generated by the DST model, utilizing the same bifurcation analysis as in the main analysis. This step further validated that the parameter values of the DST model accurately reflected the distinct craving-to-cues and cues-to-craving dynamics (**Supplementary Material 3**).

## Discussion

The main goal of this study was to contribute to understanding the multiple and complex influences of craving and cues on substance use using dynamical system analysis based on ecological momentary assessment data. To achieve this, two SARIMAX models were tested for each patient, resulting in 154 patients aligning better with *SARIMAX-model 1* (“cues-to-craving”) and 57 with *SARIMAX-model 2* (“craving-to-cues”). Parameter sets were defined through a simulation analysis to propose two DST models, representing the evolutionary and interactional dynamics of cues, craving, and use for patients corresponding to each SARIMAX model.

Figure 4 illustrates the sequence of interactions between the variables in *DST-model 1*, from which two main results could be drawn. The first finding offers insights into the mechanism of cessation of use in these patients. Specifically, cessation of use was not due to the saturation of use itself (i.e., the point where further increase is impossible due to reaching maximum capacity). Instead, cessation resulted from a specific sequence involving cue reporting and craving. In this model, a peak in cue reporting initially occurs. Once this peak is reached, we observe a gradual reduction in use, mediated by craving levels. This decrease in cue reporting returns to the baseline level, ultimately leading to cessation of use. In other words, cessation may have been triggered by the preceding peak in cue reporting, modulated by craving levels. The second finding of this model addresses the complexity of the interaction between craving and use. As observed in practice and documented in many studies, craving leads to an increase in use [2]. In this model, cue reporting precedes craving, but craving peaks simultaneously with the peak in cue reporting. A high level of cue reporting and craving is followed by a gradual decrease in cue reporting to a baseline threshold, resulting in cessation of use. The direct interaction between cue reporting and use remains unclear – specifically, whether cue reporting directly saturates use, given that cue reporting precedes use in this model. However, modulation by craving clarifies the interactions among these three variables. The model suggests that cue reporting and craving rise in tandem, leading to an increase in use. At the peak of craving, cues also reach their maximum. After this peak, craving begins to decrease, which in turn reduces cue reporting to the baseline, eventually leading to cessation of use. Importantly, use does not interrupt the craving peak (since a reduction in craving is necessary once it reaches its peak). The conclusion here is that use plays a secondary role: it follows craving, which controls the peak in cue reporting, and it also follows the reduction in cue reporting.

**Figure 4.**
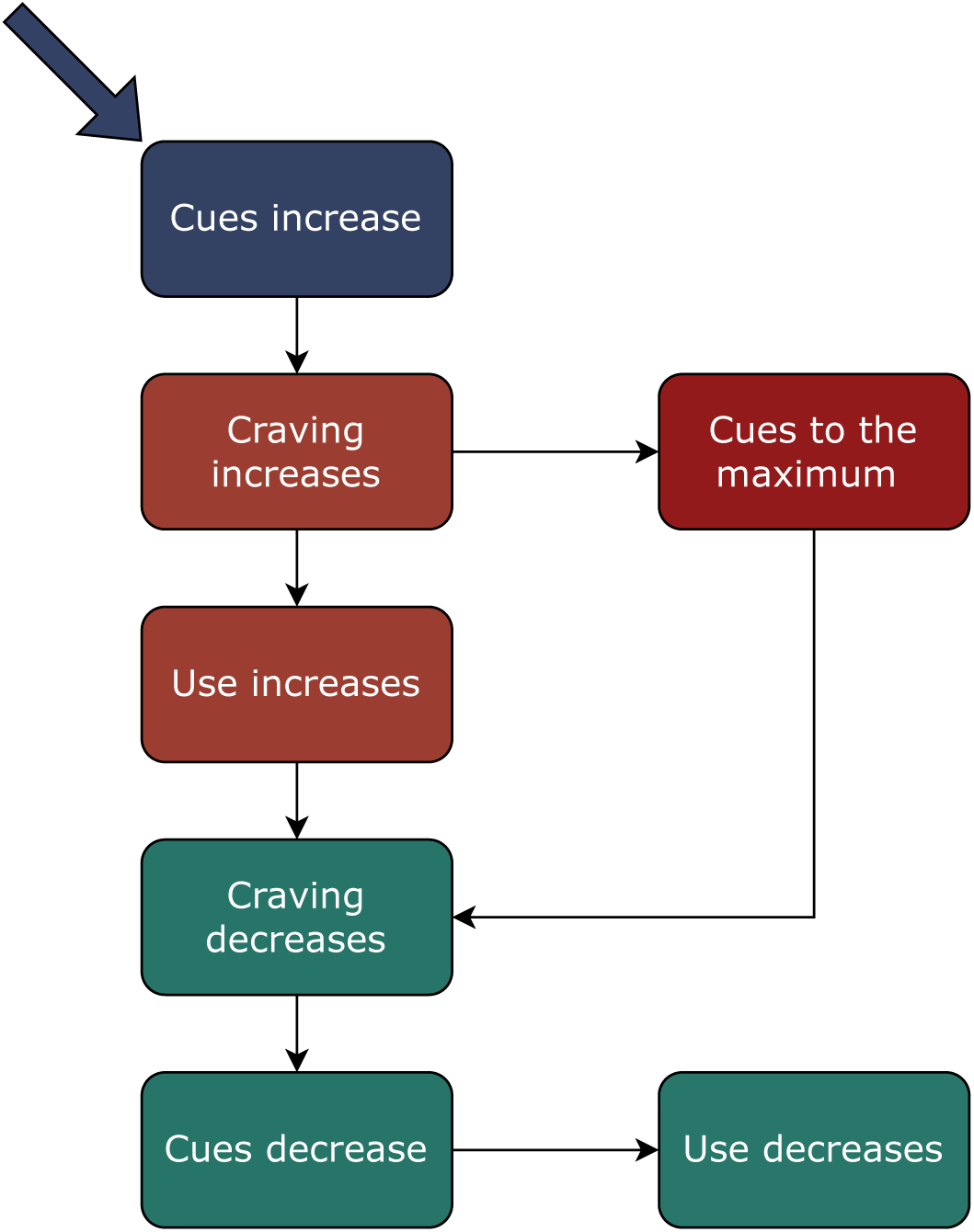
Temporal sequence of the interaction of the three variables of use, craving and cues in the *DST-model 1* (“cues-to-craving”). These relationships are described here in a linear manner in order to facilitate understanding but should be interpreted in a non-linear manner (in particular with mutual and reciprocal interactions) within the working framework of the DST. The arrow indicates the trigger variable in this model.

Figure 5 illustrates the sequence of interactions between the variables in *DST-model 2*. The primary result from this model pertains to the role of use in the relationships between craving, cues, and use. In this model, craving increases and simultaneously (or immediately after) enhances cue reporting. These reporting of cues do not exceed a relatively low threshold, resulting in a “noisy” appearance. This scattered pattern may reflect the intermittent encounter or perception of cues by patients, reflecting cue reporting, consistent with the DST-model 2, where cue reporting fluctuations do not align directly with craving peaks but rather create a baseline disturbance, testifying to a strong impact of craving on use without the presence of clearly defined prior cue reporting – consistent with recent experimental data [43]. At the same time, craving drives an increase in use. This increase is linked not only to heightened craving but also to greater cue reporting. Interestingly, the decrease in cue reporting does not drive the reduction in use or craving, as no maximum threshold of cue reporting is reached in this scenario (unlike in *DST-model 1*). Once use has peaked, it must necessarily decline. At this point, craving returns to its baseline, appearing to be influenced by the reduction in use. When use reaches maximum saturation, there is a sharp decrease in craving. The dynamics of this model may thus be as follows: initially, craving may lead to a peak in use, while simultaneously amplifying cue reporting that could be mediated by an increased perception of cues. Once use hits its peak, it may sharply reduce craving, suggesting a complex feedback loop between use and craving.

**Figure 5.**
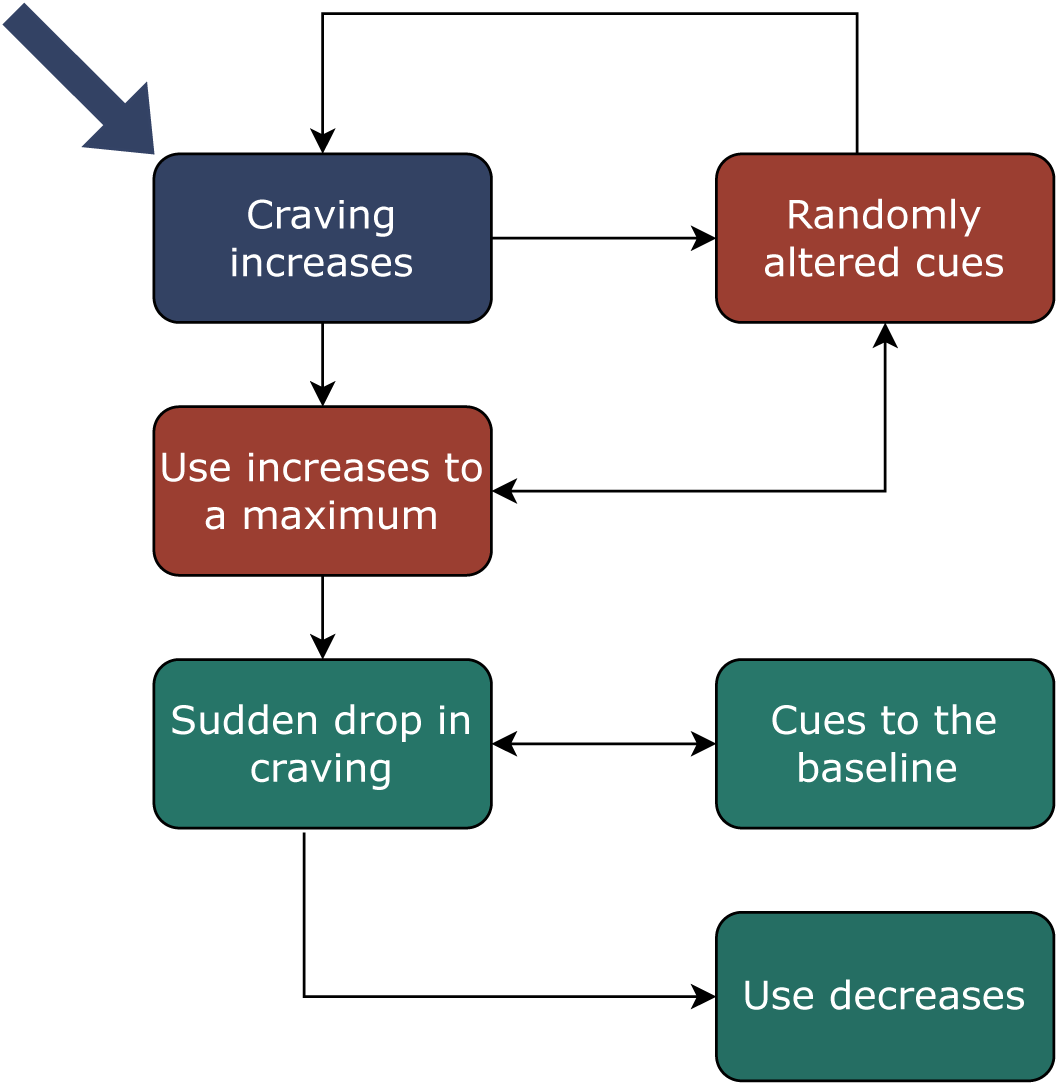
Temporal sequence of the interaction of the three variables of use, craving and cues in the *DST-model 2* (“craving-to-cues”). These relationships are described here in a linear manner in order to facilitate understanding but should be interpreted in a non-linear manner (in particular with mutual and reciprocal interactions) within the working framework of the DST. The arrow indicates the trigger variable in this model.

To summarize, when cue reporting precedes craving, the process appears to be primarily driven by a phenomenon of “maximum cue reporting saturation”, which initially increases craving and use, then leads them to decrease as cue reporting subsides. When craving precedes cue reporting, the process seems primarily driven by a phenomenon of “maximum use saturation”, which causes craving and cue reporting to decrease once use reaches its peak. In both profiles, craving serves as a critical modulator between cue reporting and use. These findings align with animal models demonstrating spontaneous exhaustion, or saturation in use [52].

The literature includes numerous simulation studies on psychiatry and DST (e.g., [15–34]); however, to our knowledge, no DST study has yet been conducted in SUD. Most existing studies focus on a single (phenotypic) variable (e.g., [31]) and have not integrated phenotype, internal factors, and environment together – three elements essential for a nuanced understanding of clinical practice. Furthermore, only eight studies have applied empirical data within DST models in psychiatry [19, 30, 53–58], and none in the context of SUD. Finally, the integration of a linear statistical method with non-linear computational analysis appears to be unprecedented.

This study’s method provides meaningful perspectives on the relationship of environmental, subjective, and behavioral variables. Such a dynamic systems approach allows for rigorous and time-sensitive modeling of clinical reasoning processes. This focus on dynamic markers aimed at reducing use is particularly novel. Additionally, based on EMA, DST could help address several other questions in the field of SUD, such as providing more detailed simulation visualizations of patient progression, studying phenotype variability, improving the understanding of dynamic attractors, structuring data from longitudinal cohorts, and potentially refining nosology with dynamic factors [59–61].

This study encompasses limitations. Firstly, the list of cues was pre-identified with the patient. However, in this study we were not trying to model the type of cues but rather change in number (quantity) of cues, and their interactions with craving. Also, what is actually measured by EMA was the reporting of cues when asked, that implies that cues were available and perceived. Only perceived cues can be reported. Whitin these data, we cannot determine if change in cue reporting was driven by change in the number of cues available to the patient, that may change with time, or by a change in state (such as craving variation), that influences the quality of cue perception, independently of cue presence. Secondly, we were trying to model rapid temporal sequences between variables, based on data that is aggregated over 4-hour periods; within these windows, it is not possible to know the mutual influence between variables. More frequent EMA measurements would provide better precision. Thirdly, at the linear level, the present study focused on analyzing the interaction between cue reporting and craving as a priority, considering that the use behavior was an outcome variable in relation to cue perception and craving. This choice raises the concern that, in linear models, the analysis of cue reporting might merely reflect cue presence that may be consequential to use patterns, and that future (SARIMAX) analyzes may integrate the use as an (exogenous) influence variable. Moreover, certain statistical choices were necessarily made for these linear models (e.g., favoring AIC over BIC), methods that could be compared through simulations, for instance. However, beyond this potential extension to linear models, the use of DST models allows us to highlight the interactions between use, craving, and cues. Fourthly, although subgroup analyses by SUD type (alcohol, tobacco, cannabis, opiates, and cocaine) could provide valuable insights, incorporating them could overcomplicate the already substantial findings. Models were developed by combining different substance groups based on evidence that cues influence craving, and craving, in turn, affects substance use and relapse across various types of SUD [2–4]. Fifthly, a known limitation of the SARIMAX models is that they evaluate each patient as aligning with one model, though a patient’s condition may not be stable over time, and may change from one model to the other, and there is the possibility that the two models could co-exist for some individuals. Furthermore, SARIMAX models have difficulty handling nonlinear relationships and complex data patterns, requiring significant fine-tuning to achieve optimal performance and assuming stationarity in the data. We sought to address these limitations by verifying stationarity, performing a grid search to optimize the models, and subsequently employing nonlinear models for greater precision. In parallel, the nonlinear models based on DST also present certain methodological constraints [28]. For instance, it could be necessary to refine the model to modelize symptoms and behaviors of different types (and not only “use” or “craving”), different set of parameters or differential equations could have achieved relatively similar dynamics and the qualitative interpretation of nonlinear phenomena, although constrained by clinical perspectives, remains challenging. Different parameter sets could yield similar data fits, highlighting the critical need for robust model validation.

### Conclusion

In conclusion, this study introduces an innovative approach that combines EMA data, temporal statistical models (SARIMAX), and computational models (DST) to examine the nonlinear and interconnected relationships between cues, craving, and use in substance use disorders (SUD). The results could have important implications for the temporal phenotyping of patients, helping to refine the complex patterns of influence among SUD variables and supporting the identification of temporally anchored markers and mechanisms in clinical practice – thus opening critical therapeutic avenues for precision addiction treatments. Moving forward, validating the robustness of the model will remain essential, further reinforcing the reliability of these findings and their potential to advance tailored approaches in addiction treatment.

## Acknowledgements

The authors express theirs thanks to all participants for their contribution and are grateful to all the interviewers of the Addiction Team of SANPSY lab 2009-2022.

## Data availability

The data that support the findings of the current manuscript are available from the corresponding author, upon reasonable request.

## Authors’ contributions

Marc Auriacombe and Fuschia Serre were the principal investigators, obtaining funding and supervised the study. Marc Auriacombe provided access to participants. Damien Depannemaecker and Christophe Gauld conceptualized and undertook analysis, interpretation of data and drafting of manuscript. Fuschia Serre participated in patient recruitment, data collection and database management. Marc Auriacombe and Fuschia Serre sustained the elaboration of the methodology and writing. All authors undertook the critical revision of the manuscript for important intellectual content and all authors significantly contributed to the manuscript and approved the final version.

## Footnotes

1 *x*: symptom intensity, representing use; *y*: participant’s internal factors, representing craving intensity; *z*: environmental noise, representing the reporting of cues. τ***_x,y,z,f_***: time scales of the four equations; S_ₘₐₓ_: maximal symptom level; R_ₛ_ and R_ᵦ_: sensitivity, with R_ₛ_ for symptom sensitivity and R_ᵦ_ for internal element (potentiation) sensitivity; λ_ₛ_ and λ_ᵦ_: slope of symptom/internal element curves, with λ_ₛ_ affecting symptom intensity based on patient state γ, and λ_ᵦ_ affecting internal elements based on environment z; P: maximal rate of internal elements, aggregating biological, psychological, and phenomenological factors, shaping phenotype from biological signals; L: level of predisposing factors contributing to a permanent shift in internal elements; λ***_f_***: scaling factor for slow fluctuations affecting L; S: overall environmental sensitivity; α and β: weights of x and y effects on environmental perception.

## References

1. Auriacombe M, Serre F, Denis C, Fatseas M. Diagnosis of addictions. Chapter 11. The Routledge Handbook of the Philosophy and Science of Addiction 2018. p. 132–144.

2. Vafaie N, Kober H. Association of Drug Cues and Craving With Drug Use and Relapse: A Systematic Review and Meta-analysis. JAMA Psychiatry. 2022;79:641–650.

3. Serre F, Fatseas M, Denis C, Swendsen J, Auriacombe M. Predictors of craving and substance use among patients with alcohol, tobacco, cannabis or opiate addictions: Commonalities and specificities across substances. Addict Behav. 2018;83:123–129.

4. Serre F, Fatseas M, Swendsen J, Auriacombe M. Ecological momentary assessment in the investigation of craving and substance use in daily life: a systematic review. Drug Alcohol Depend. 2015;148:1–20.

5. Cavicchioli M, Vassena G, Movalli M, Maffei C. Is craving a risk factor for substance use among treatment-seeking individuals with alcohol and other drugs use disorders? A meta-analytic review. Drug Alcohol Depend. 2020;212:108002.

6. Cleveland HH, Knapp KS, Brick TR, Russell MA, Gajos JM, Bunce SC. Effectiveness and Utility of Mobile Device Assessment of Subjective Craving during Residential Opioid Dependence Treatment. Subst Use Misuse. 2021;56:1284–1294.

7. Bujarski S, Roche DJO, Sheets ES, Krull JL, Guzman I, Ray LA. Modeling naturalistic craving, withdrawal, and affect during early nicotine abstinence: a pilot EMA study. Exp Clin Psychopharmacol. 2015;23:81–89.

8. Sayette MA. The Role of Craving in Substance Use Disorders: Theoretical and Methodological Issues. Annu Rev Clin Psychol. 2016;12:407–433.

9. Stone AA, Shiffman S. Ecological Momentary Assessment (Ema) in Behavioral Medicine. Annals of Behavioral Medicine. 1994;16:199–202.

10. Serre F, Fatseas M, Debrabant R, Alexandre J-M, Auriacombe M, Swendsen J. Ecological momentary assessment in alcohol, tobacco, cannabis and opiate dependence: a comparison of feasibility and validity. Drug Alcohol Depend. 2012;126:118–123.

11. Fatseas M, Serre F, Alexandre J-M, Debrabant R, Auriacombe M, Swendsen J. Craving and substance use among patients with alcohol, tobacco, cannabis or heroin addiction: a comparison of substance- and person-specific cues. Addiction. 2015;110:1035–1042.

12. Ellis JD, Mun CJ, Epstein DH, Phillips KA, Finan PH, Preston KL. Intra-individual variability and stability of affect and craving among individuals receiving medication treatment for opioid use disorder. Neuropsychopharmacology. 2022;47:1836–1843.

13. Burgess-Hull AJ, Panlilio LV, Preston KL, Epstein DH. Trajectories of craving during medication-assisted treatment for opioid-use disorder: Subtyping for early identification of higher risk. Drug Alcohol Depend. 2022;233:109362.

14. Panlilio LV, Stull SW, Kowalczyk WJ, Phillips KA, Schroeder JR, Bertz JW, et al. Stress, craving and mood as predictors of early dropout from opioid agonist therapy. Drug Alcohol Depend. 2019;202:200–208.

15. Kelso JAS. Dynamic patterns: The self-organization of brain and behavior. Cambridge, MA, US: The MIT Press; 1995.

16. Boldrini M, Placidi G, Marazziti D. Applications of Chaos Theories to Psychiatry: A Review and Future Perspectives. CNS Spectrums. 1998;3:22–29.

17. Ayers S. The Application of Chaos Theory to Psychology. Theory & Psychology. 1997;7:373–398.

18. Barton S. Chaos, self-organization, and psychology. Am Psychol. 1994;49:5–14.

19. Bonsall MB, Geddes JR, Goodwin GM, Holmes EA. Bipolar disorder dynamics: affective instabilities, relaxation oscillations and noise. J R Soc Interface. 2015;12:20150670.

20. Bystritsky A, Nierenberg AA, Feusner JD, Rabinovich M. Computational non-linear dynamical psychiatry: a new methodological paradigm for diagnosis and course of illness. J Psychiatr Res. 2012;46:428–435.

21. Chang S-S, Chou T. A Dynamical Bifurcation Model of Bipolar Disorder Based on Learned Expectation and Asymmetry in Mood Sensitivity. Computational Psychiatry. 2018;2:205–222.

22. Cramer AOJ, Van Der Sluis S, Noordhof A, Wichers M, Geschwind N, Aggen SH, et al. Dimensions of Normal Personality as Networks in Search of Equilibrium: You Can’t like Parties if you Don’t like People. Eur J Pers. 2012;26:414–431.

23. Cramer, Borkulo CD van, Giltay EJ, Maas HLJ van der, Kendler KS, Scheffer M, et al. Major Depression as a Complex Dynamic System. PLOS ONE. 2016;11:e0167490.

24. Cui J, Lichtwarck-Aschoff A, Olthof M, Li T, Hasselman F. From Metaphor to Computation: Constructing the Potential Landscape for Multivariate Psychological Formal Models. Multivariate Behavioral Research. 2023;58:743–761.

25. Cui J, Hasselman F, Lichtwarck-Aschoff A. Unlocking nonlinear dynamics and multistability from intensive longitudinal data: A novel method. 2023. 1 December 2023.

26. Demic S, Cheng S. Modeling the Dynamics of Disease States in Depression. PLoS One. 2014;9:e110358.

27. Durstewitz D, Huys QJM, Koppe G. Psychiatric Illnesses as Disorders of Network Dynamics. Biol Psychiatry Cogn Neurosci Neuroimaging. 2021;6:865–876.

28. Gauld C, Depannemaecker D. Dynamical systems in computational psychiatry: A toy-model to apprehend the dynamics of psychiatric symptoms. Frontiers in Psychology. 2023;14.

29. Globus GG, Arpaia JP. Psychiatry and the new dynamics. Biological Psychiatry. 1994;35:352–364.

30. Gottschalk A, Bauer MS, Whybrow PC. Evidence of Chaotic Mood Variation in Bipolar Disorder. Archives of General Psychiatry. 1995;52:947–959.

31. Haken H, Tschacher W. How to Modify Psychopathological States? Hypotheses Based on Complex Systems Theory. Nonlinear Dynamics Psychology and Life Sciences. 2017;21:19–34.

32. Haslbeck J, Ryan O, Robinaugh D, Waldorp L, Borsboom D. Modeling Psychopathology: From Data Models to Formal Theories. 2019. December 2019. None.

33. Hayes AM, Yasinski C, Barnes JB, Bockting CLH. Network destabilization and transition in depression: New methods for studying the dynamics of therapeutic change. Clin Psychol Rev. 2015;41:27–39.

34. Hayes AM, Laurenceau J-P, Feldman G, Strauss JL, Cardaciotto L. Change is Not Always Linear: The Study of Nonlinear and Discontinuous Patterns of Change in Psychotherapy. Clin Psychol Rev. 2007;27:715–723.

35. Witkiewitz K, Marlatt GA. Modeling the complexity of post-treatment drinking: it’s a rocky road to relapse. Clin Psychol Rev. 2007;27:724–738.

36. Foo JC, Noori HR, Yamaguchi I, Vengeliene V, Cosa-Linan A, Nakamura T, et al. Dynamical state transitions into addictive behaviour and their early-warning signals. Proceedings of the Royal Society B: Biological Sciences. 2017;284:20170882.

37. Serre F, Fatseas M, Swendsen J, Auriacombe M. Does craving intensity influence cue exposure reports? An ecological momentary assessment study in patients with alcohol, tobacco, cannabis and heroin use disorder. Drug and Alcohol Dependence. 2015;C:e201.

38. Fatseas M, Serre F, Swendsen J, Auriacombe M. Effects of anxiety and mood disorders on craving and substance use among patients with substance use disorder: An ecological momentary assessment study. Drug Alcohol Depend. 2018;187:242–248.

39. Denis C, Fatséas M, Beltran V, Serre F, Alexandre J-M, Debrabant R, et al. Usefulness and validity of the modified Addiction Severity Index: A focus on alcohol, drugs, tobacco, and gambling. Subst Abus. 2016;37:168–175.

40. McLellan AT, Kushner H, Metzger D, Peters R, Smith I, Grissom G, et al. The Fifth Edition of the Addiction Severity Index. J Subst Abuse Treat. 1992;9:199–213.

41. Sheehan DV, Lecrubier Y, Sheehan KH, Amorim P, Janavs J, Weiller E, et al. The Mini-International Neuropsychiatric Interview (M.I.N.I.): the development and validation of a structured diagnostic psychiatric interview for DSM-IV and ICD-10. J Clin Psychiatry. 1998;59 Suppl 20:22-33;quiz 34-57.

42. Fatseas M, Serre F, Swendsen J, Auriacombe M. Effects of anxiety and mood disorders on craving and substance use among patients with substance use disorder: An ecological momentary assessment study. Drug and Alcohol Dependence. 2018;187:242–248.

43. Serre F, Gauld C, Lambert L, Baillet E, Beltran V, Daulouede J, et al. Predictors of substance use during treatment for addiction: A network analysis of ecological momentary assessment data. Addiction. 2024:add.16658.

44. Chirokoff V, Dupuy M, Abdallah M, Fatseas M, Serre F, Auriacombe M, et al. Craving dynamics and related cerebral substrates predict timing of use in alcohol, tobacco, and cannabis use disorders. Addiction Neuroscience. 2023;9:100138.

45. Shumway RH, Stoffer DS. ARIMA Models. In: Shumway RH, Stoffer DS, editors. Time Series Analysis and Its Applications: With R Examples, Cham: Springer International Publishing; 2017. p. 75–163.

46. Mushtaq R. Augmented Dickey Fuller Test. 2011.

47. Vagropoulos SI, Chouliaras GI, Kardakos EG, Simoglou CK, Bakirtzis AG. Comparison of SARIMAX, SARIMA, modified SARIMA and ANN-based models for short-term PV generation forecasting. 2016 IEEE International Energy Conference (ENERGYCON), 2016. p. 1–6.

48. Alharbi FR, Csala D. A Seasonal Autoregressive Integrated Moving Average with Exogenous Factors (SARIMAX) Forecasting Model-Based Time Series Approach. Inventions. 2022;7:94.

49. Strogatz SH. Nonlinear Dynamics And Chaos: With Applications To Physics, Biology, Chemistry, And Engineering. 1st edition. Cambridge, Mass: CRC Press; 2000.

50. Izhikevich EMM. Dynamical Systems in Neuroscience: The Geometry of Excitability and Bursting. 2438 3rd edition. Cambridge, Massachusetts London, England: The MIT Press; 2010.

51. Van Rossum G, Drake FL. Python 3 Reference Manual. Scotts Valley, CA: CreateSpace; 2009.

52. Ahmed SH. Validation crisis in animal models of drug addiction: Beyond non-disordered drug use toward drug addiction. Neuroscience & Biobehavioral Reviews. 2010;35:172–184.

53. Pezard L, Nandrino JL, Renault B, ElMassioui F, Allilaire JF, Muller J, et al. Depression as a dynamical disease. Biol Psychiatry. 1996;39:991–999.

54. Tschacher W, Scheier C, Hashimoto Y. Dynamical analysis of schizophrenia courses. Biological Psychiatry. 1997;41:428–437.

55. van de Leemput IA, Wichers M, Cramer AOJ, Borsboom D, Tuerlinckx F, Kuppens P, et al. Critical slowing down as early warning for the onset and termination of depression. Proceedings of the National Academy of Sciences. 2014;111:87–92.

56. Tschacher W, Haken H. Causation and chance: Detection of deterministic and stochastic ingredients in psychotherapy processes. Psychotherapy Research. 2020;30:1075–1087.

57. Odgers CL, Mulvey EP, Skeem JL, Gardner W, Lidz CW, Schubert C. Capturing the ebb and flow of psychiatric symptoms with dynamical systems models. Am J Psychiatry. 2009;166:575–582.

58. Heiby E, Pagano I, Blaine D, Nelson K, Heath R. Modeling Unipolar Depression as a Chaotic Process. Psychological Assessment. 2003;15:426–434.

59. Wichers M, Schreuder MJ, Goekoop R, Groen RN. Can we predict the direction of sudden shifts in symptoms? Transdiagnostic implications from a complex systems perspective on psychopathology. Psychol Med. 2019;49:380–387.

60. Nelson B, McGorry PD, Wichers M, Wigman JTW, Hartmann JA. Moving From Static to Dynamic Models of the Onset of Mental Disorder: A Review. JAMA Psychiatry. 2017;74:528–534.

61. Salvi JD, Rauch SL, Baker JT. Behavior as Physiology: How Dynamical-Systems Theory Could Advance Psychiatry. AJP. 2021;178:791–792.

